# The Effect of Regionalization of Thoracic Surgery on Blood Transfusion at the Ottawa Hospital

**DOI:** 10.64898/2026.10.15.26365639

**Authors:** Jacob Smith, Benjamin Smith, Tyler Pittman

## Abstract

This paper examines the impact of Cancer Care Ontario’s regionalization of lung cancer surgery on blood transfusion use for thoracic surgery at The Ottawa Hospital. Using aggregate data scraped from the paper “Changing trends in blood transfusion: an analysis of 244,013 hospitalizations” by Shehata et al. (2014) we estimated the effect regionalization on the log mean number of blood transfusions among thoracic surgeons (who treat lung cancer), utilizing other surgical specialties as controls. We found that the average number of blood transfusions increased significantly among thoracic surgeons following the opening of a new cancer care centre at the hospital in December 2009, while control specialties showed no parallel change. Contextualizing these results in a modified “population level” treatment threshold model, it is argued that the main driver of this increase in blood transfusions among thoracic surgeons was due to the influx of less skilled thoracic surgeons to The Ottawa Hospital, and the closing of thoracic surgery units at smaller hospitals which was part of the broader regionalization process.

## 1. Introduction

Blood, plasma, and other blood-based products are vital inputs for emergency treatments, trauma care, and scheduled surgeries. Given their clinical importance and scarcity, existing literature has primarily investigated supply-side incentives for donation and clinical guidelines governing transfusion thresholds (Sun, Lu, & Jin, 2016; Callum et al., 2023). However, empirical evidence on provider-level determinants of blood utilization and the role of institutional reorganization remains sparse. In Canada, blood resource management is centralized through Canadian Blood Services, which allocates inventories to hospitals (Gabarin & Yan, 2024). Once allocated at the hospital level, the final utilization decisions rest entirely with individual physicians, who guided by clinical discretion and patient health status. While professional guidelines, such as the Frankfurt consensus adopted by The Ottawa Hospital, recommend red blood cell transfusion when hemoglobin concentrations fall below 7.5 g/dL (Mueller et al., 2019; OHRI, 2019; Mazer et al., 2017), documented evidence reveals persistent practice-style variation and inappropriate utilization across Ontario hospitals (Fang et al., 2025; Lin et al., 2024; Spradbrow et al., 2016). Because blood transfusions are not billed as separate line items within the provincial schedule of medical benefits, treatment intensity is governed strictly by physician judgment rather than explicit fee incentives. Consequently, these patterns reflect differences in physician beliefs, risk tolerance, and clinical judgment. Using this setting, this paper examines how Cancer Care Ontario’s regionalization of lung cancer surgery affected blood transfusion rates in thoracic surgery at The Ottawa Hospital. From the evaluation of a clinical decision-making framework, we argue that the main driver of changes in blood use is the integration of surgeons from smaller community hospitals who hold different practice styles and clinical views.

This paper contributes to the literature evaluating the regionalization of surgical care. A well-known finding in cancer care from Ontario, Canada concluded that regionalization did not impact operative mortality and increased procedure volumes (Bendzsak A. M., Baxter, Darling, Austin, & Urbach, 2017). Other research on the effect of the regionalization of cancer care in Ontario focused on other patient outcomes and institutional objectives with general the policy ex-post (Nason, et al., 2020; Grilli, Violi, Bassi, &Marino, 2021). Previous analysis of this policy change has largely overlooked how the utilization of secondary hospital resources—specifically blood products—changed following this centralization of thoracic surgery. While existing evaluations focused primarily on mortality and assessing wait times, this paper is the first to examine the unintended consequences of regionalization on blood utilization. This paper also contributes to the literature on medical decision-making and treatment thresholds. The mechanism driving changes in blood utilization stems from the institutional consolidation of thoracic surgeons moving from 46 sites into 14 designated regional centers. Because blood transfusions carry no explicit financial incentives or unbundled fee-for-service codes, transfusion decisions are driven entirely by a physician’s subjective assessment of the patient benefits from blood transfusion. We frame this behavior using the canonical “Treatment Threshold Model” introduced by Paucker and Kassirer (1975), which has been extensively validated in theoretical and simulated environments (Djulbegovic et al., 2014; Felder & Mayerhofer, 2021). This paper uniquely characterizes changes in hospital-level blood utilization through a population analogue of the Pauker-Kassirer framework, demonstrating how shifting distributions of physician *treatment thresholds* for blood transfusion, which in this case is brought on through change in composition of surgeons at The Ottawa Hospital, alter aggregate blood resource use.

The remainder of this paper will proceed as follows: Section 2 presents the institutional background on the regionalization of lung cancer care in Ontario. Sections 3 and 4 present the primary data sources with descriptive statistics and characterize a modified threshold model which formalizes our hypotheses. Section 5 presents the empirical strategy, including identifying assumptions and proposed robustness tests. Sections 6 and 7 present the main results, robustness tests and discuss these results and limitations. Section 8 concludes.

## 2. Institutional Background and Clinical Environment

### 2.1 The Regionalization of Cancer Care in Ontario

Historically, Cancer Care Ontario (CCO) is an organization which existed independently before being formally integrated into the Ontario Ministry of Health. It was first established by name in 1997 but existed as Ontario Cancer Treatment Research Foundation since 1943. A defining policy milestone of this era occurred in April 2007, when CCO enacted a major policy to regionalize complex cancer surgeries starting most notably with lung cancer surgery limiting procedures to specifically designated high-volume hospitals, reducing the number of thoracic surgery units from 46 to 14 centers in Ontario (Sundaresan, et al., 2013). This was in part based on evidence from Dudley et al. (2000), which found improved outcomes in patients treated in higher volume hospitals. This involved the ongoing redirection of patients, thoracic surgeons and thoracic anesthesiologists from lower volume hospitals to larger volume hospitals.

The regionalization of cancer care in Ontario began formally in April 2007, however, this did not impact The Ottawa Hospital until December 2009 when their cancer centre was opened to accommodate these proposed changes (Infrastructure Ontario, 2010). Note, regionalization did not change the surgeon to which the patients were assigned, but rather only changed the location where procedures took place. Thus, the only change which occurred was the movement of patients, thoracic surgeons and other personnel from smaller thoracic surgery units to The Ottawa Hospital. From the perspective of this hospital system, there was a simultaneous expected increase in both surgical capacity and manpower. The fact that patient assignment to surgeons remains fixed simplifies characterization fluctuations in blood use and will be explained in the next section.

## 3. Theoretical Framework

### 3.1 The Classic Treatment Threshold Model

The classic treatment threshold model considers a physician faced with a binary decision to treat or not treat a patient of uncertain health status—specifically, whether a patient determined likely to be sick enough to warrant an otherwise risky or costly intervention. The version of this model presented in this section is based on the presentation from Felder and Mayrhofer (2021). In this model a physician makes a choice between two expected utility equations *EU*_*T*_ which denotes the expected utility from treatment and *EU*_*NT*_ which denotes the expected utility of no treatment. The physician is assumed to be entirely benevolent and thus only values the payoffs based on patient outcomes from each health state defined by *u*(⋅). Formally, the physician’s problem is defined as:

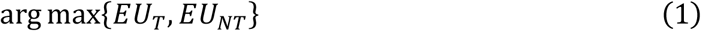

where:

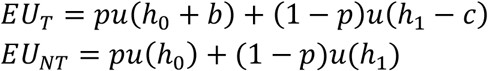

Here, *p* is the physician’s prior belief that the patient is in the worse health state *h*_0_, *b* is the clinical benefit of treatment when sick, and *c* is the harm or cost of treatment if administered to a healthy patient. The underlying health states and parameters satisfy the standard ranking:

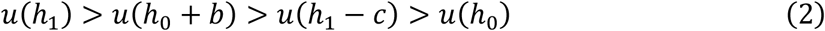

The treatment threshold, 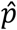, is defined as probability the physician is indifferent between treating and not treating (*EU*_*T*_ = *EU*_*NT*_). This probability can be thought of as the minimal probability for the sick state required for the physician to administer treatment. Solving for 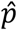 yields

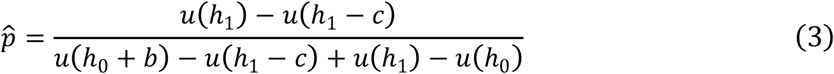

In this framework, the individual threshold depends on the net utility gains and losses associated with treatment. To make sense of this model in a clinical context, a conceptual model of this model is displayed in Figure 1. A patient seeking treatment first arrives, the attending physician then forms a prior *p* and compares it against the threshold 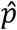. Treatment is provided if and only if 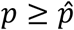 Note that this model does not speak to how physicians form their beliefs. Patients may arrive with an assigned probability of being sick which is only evaluated by the physician directly. This model defines how treatment decisions may be formed based on patient outcomes alone in a two-arm decision problem.

**Figure 1:**
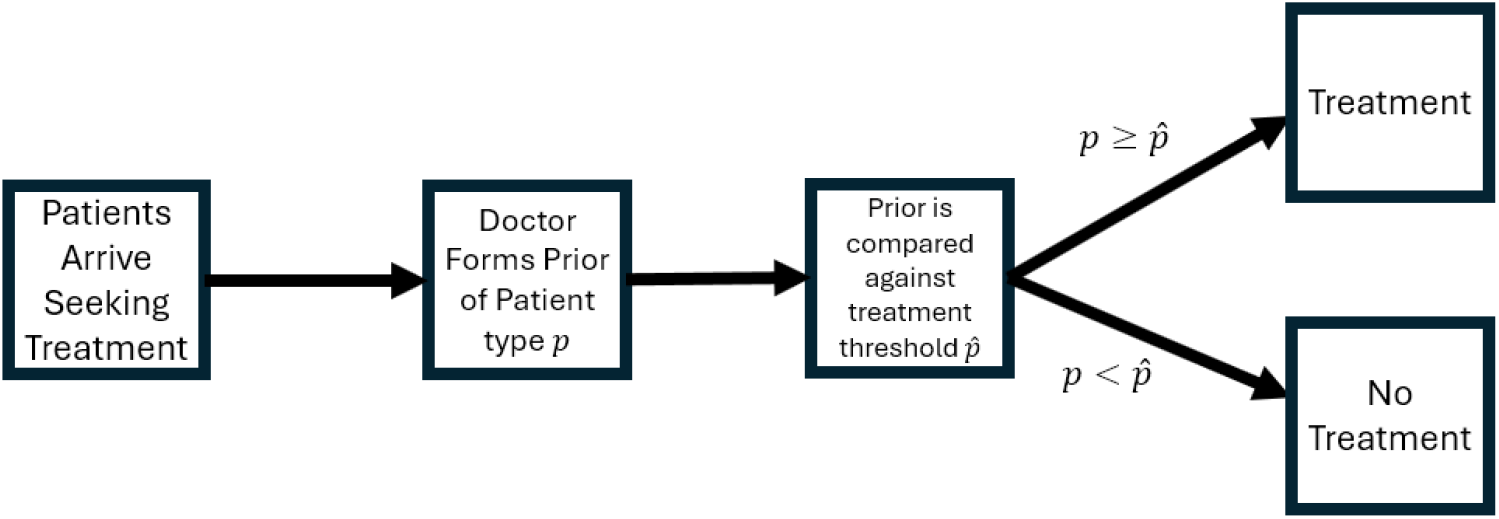
The Physician’s treatment decision conceptualized graphically.

### 3.2 Adapting the Model to a Population Context

To bridge this individual decision rule to population-level outcomes which model hospital resource use, we allow the underlying clinical success and failure rates to follow distributions that preserve an expected utility ranking through first-order stochastic dominance. This ranking is initially set through some unobserved previous history *θ* up to this time. Appealing to the derivation of the treatment threshold directly, the hospital level treatment threshold 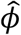 is

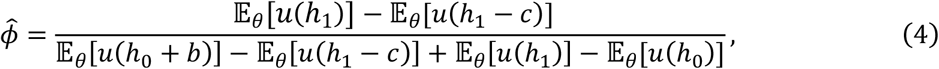

where

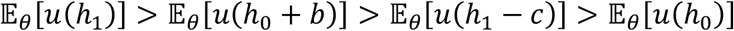

and

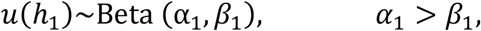

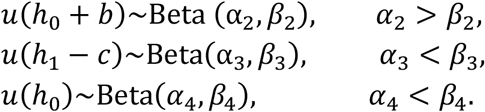

Explicitly, we model the valuation of clinical outcomes valued by utility functions using Beta distributions which are parameterized by shape parameters *α* and *β*. Beta distributions are a natural fit for this context because their shape parameters map directly to binary patient outcome data (such as survival versus mortality counts). For instance, if *n* patients are treated with *s* successes and *f* failures, the conjugate Beta distribution updates with parameters corresponding to these clinical counts. If we are interested in determining the distribution of the utility of *u*(*h*_0_ + *b*)we simply count the number of patients with this outcome from the total number of patients who receive treatment. If *s* is the total number of patients with this successful treatment outcome, then *α* = *s* and *β* = *n− s*. If we are modelling adverse outcomes we would set *β* as the number of “successes” (i.e., number of patients who fall into this category) and alpha would be the remaining set of patients. To illustrate if we are interested in identifying the distribution for *u*(*h*_0_), *β* would correspond to the number of patients in this state while *α* would correspond to otherwise healthy patients.

We can apply this framework to evaluate regionalization policies—such as the consolidation of thoracic surgery from smaller community hospitals to larger regional centers. Given that smaller hospitals exhibit higher post-operative complication and mortality rates (Silber, et al., 2010; Fareed, 2012; Chen, Duncan, Talarico, McIsaac, & McIsaac, 2024), the difference between smaller (*S*) and larger (*L*) hospitals, respectively, is such that:

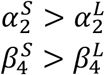

Note, that *α* and *β* in this context are not abstract shape parameters, but rather correspond to the number of patient types seen in each hospital. This calibrates the distribution of outcomes directly. When regionalization occurred, there was a movement of patient types at the hospital level, revising the parameter inputs of *α* and *β* directly at larger hospitals. This is not the addition of distributions. Rather, it revises the input parameters characterizing the Beta distributions of the underlying hospital threshold rule. This may be thought of as a revision of the “collective memory” at the hospital level.

Mechanically, the mean of a Beta distribution is given by 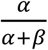 Thus, higher adverse outcome parameters reduce the treatment threshold. This core mechanism generates testable comparative statics that we bring directly to the administrative panel dataset. Specifically, that following the regionalization of thoracic surgery, the propensity to treat for blood transfusions at The Ottawa Hospital increased due to the increase in smaller hospital surgeons who have lower treatment thresholds.

## 4. Data and Descriptive Statistics

The data for this study was obtained from Figure 5 in Shehata et al. (2014), which illustrates average red blood cell (RBC) unit usage at The Ottawa Hospital across four surgical specialties. The surgical specialties are cardiac surgery, thoracic surgery, general surgery and urology. Because the raw dataset was unavailable, values were scraped from this figure using the R package juicr (Lajeunesse, 2021). Use of this package for the figure-based extraction of study data has been suggested by Ivimey-Cook et al. (2023). This extraction yielded 272 total observations—68 per surgical specialty—spanning the timeframe from November 2006 to June 2012. Among these services, thoracic surgery exhibited the highest temporal volatility, whereas general surgery showed the least. Figure 2 presents a replication of Figure 5 Shehata et al. (2014), while Table 1 summarizes the means and standard deviations for each specialty.

**Table 1:** Descriptive Statistics of RBC use by Specialty based on scraped data.

| **Specialty** | **Mean RBC Units** | **Standard Deviation** | **Observations** |
| --- | --- | --- | --- |
| **Cardiac Surgery** | **4.732** | **1.271** | **68** |
| **Thoracic Surgery** | **3.882** | **2.426** | **68** |
| **General Surgery** | **3.657** | **0.919** | **68** |
| **Urology** | **4.587** | **1.121** | **68** |

**Figure 2:**
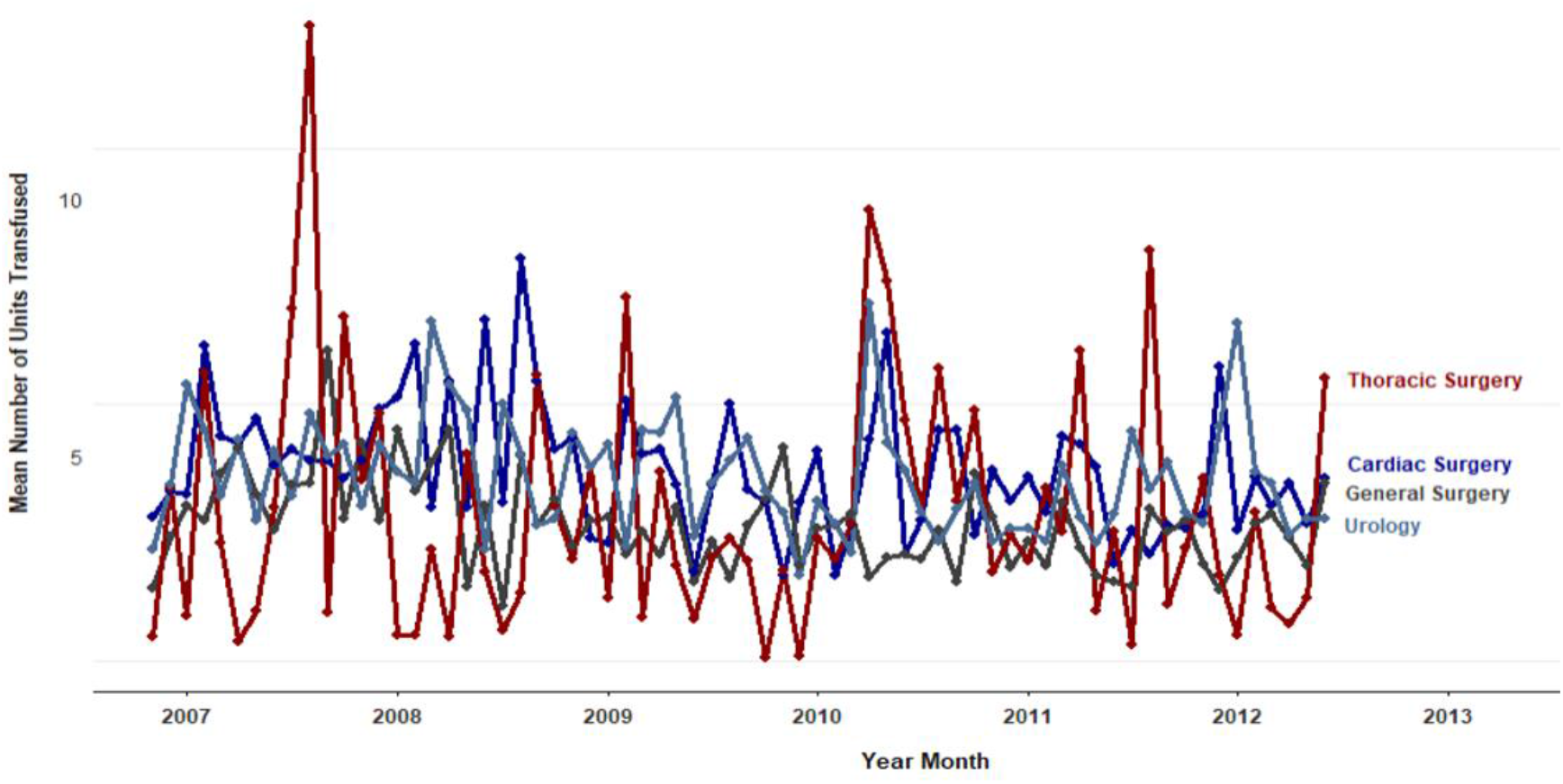
Mean Red Blood Cell Unit use by Admitting Surgical Specialty Based on Scraped Data from Figure 5 in Shehata et al (2014)

Another crucial data component involves tracking the movement of thoracic surgeons from smaller community hospitals to larger provincial centers. Since direct migration data for surgeons over this period is unavailable, we use a proxy from the Canadian Institute for Health Information’s (CIHI) National Physician Database. With this database, we obtained the total major procedure volumes and personnel counts for each specialty and computed the province-level average major procedure volumes per surgeon, displayed in Figure 3. Key trends reveal that between 2009– 2010 and 2010–2011, the average surgical volumes for thoracic and cardiovascular surgery fell by approximately 9%, dropping from 264 to 240 procedures annually. Because most thoracic surgeries were concentrated at larger hospitals, this relocation of surgeons functioned as a labor supply shock that reduced average output. In contrast, general surgery experienced a large decline in average surgical volumes in 2008 due to broader, province-wide hospital restructuring unrelated to the Cancer Care Ontario initiative (Ontario Health Coalition, 2008). Urology was the only specialty unaffected by either the 2008 restructuring or regionalization in terms of personnel, resulting in the most stable data series throughout this period. While data at this level does not allow for formal statistical testing, as there are only six observations per group, it provides descriptive support for the idea that regionalization acted as a labor supply shock to larger hospitals.

**Figure 3:**
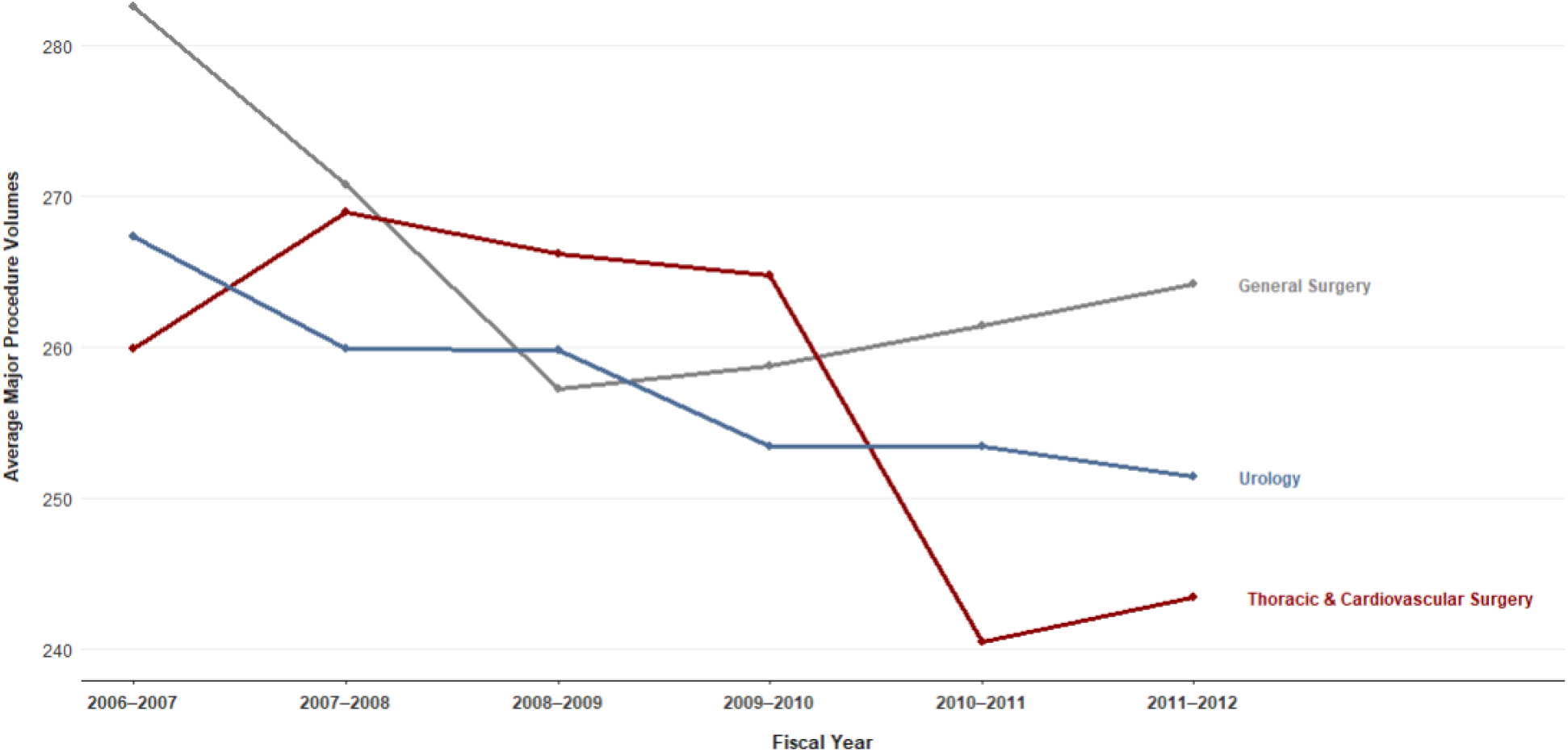
Average Major Surgery Volumes Across Surgical Specialty. 2006-2012

## 5. Empirical Strategy

### 5.1 Identification

To identify the causal effect of thoracic surgery regionalization on blood transfusion rates across specialties, we estimate a dynamic difference-in-differences model grounded in three core identifying assumptions:

1. The exogeneity and targeted nature of the regionalization policy.
2. The comparability of blood transfusion practices across surgical specialties.
3. Parallel trends in mean blood use in absence of opening of the new cancer care centre at The Ottawa Hospital.

First, we assume the policy change is exogenous to individual provider behavior. Because Ontario’s Ministry of Health directly regulates hospital organization, thoracic surgeons could neither contest nor strategically shape the restructuring. The targeted, policy-driven nature of this regionalization is further corroborated by the sharp decline in average major procedure volumes in 2010-2011 as previously documented in Figure 3.

The second and third identifying assumptions are stronger. To defend the comparability of blood use across specialties, we note that the decision to transfuse is governed by attending physicians operating under standardized clinical guidelines. Although underlying patient populations differ across specialties, the core decision-making process regarding blood administration remains independent of the regionalization shock itself. This is a decision which is made entirely at the physician level. The parallel trends assumption is fundamentally untestable; however, we support it directly through a dynamic event-study specification by examining the coefficients pre 2009 and 2010. This is done by specifying a model which interacts year indicators with specialty indicators. This way we can formally evaluate whether differential trends in blood transfusion existed across specialties prior to the opening of the new cancer center in 2009. This is expounded upon further in the subsequent section.

### 5.2 Empirical specification

To analyze the effect of regionalization and the opening of a new cancer centre at The Ottawa Hospital on blood transfusion utilization for thoracic surgery, the following dynamic differences in differences model is estimated.

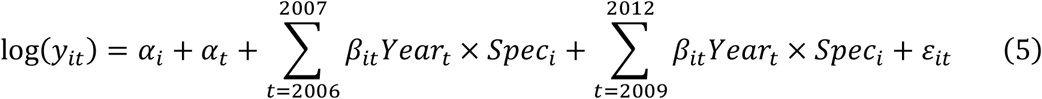

Where log(*y*_*it*_)is the natural log of the mean number of blood transfusions by specialty *i* in year *t*, *α*_*i*_ are specialty fixed effects and *α*_*t*_ are year fixed effects. The interaction terms *β*_*it*_ estimate the specialty specific deviations from 2008, which is the year before the regionalization impacted The Ottawa Hospital and the opening of its new cancer centre. Each *β*_*it*_ in this context may be interpreted as a semi elasticity of mean blood transfusion utilization with respect to procedure volumes. This framework allows for the testing of any resulting change from the decline in procedure volumes. Throughout, the identifying assumption is that any change in transfusion utilization would be brought on by a change in average procedure volumes by specialty. Further, it is assumed that during this period, no change to blood use protocols occurred at the hospital level. The assumption about the targeted nature of change in cardiovascular and thoracic surgery procedures will be inspected in the context of the event study plots. To further assess the robustness of these results, we estimate the effect of the opening of the cancer center in December 2009 on mean blood use in cardiovascular surgery and urology using general surgery use as a control.

## 6. Results

The regionalization of lung cancer surgery in Ontario and the opening of the new cancer center at The Ottawa Hospital resulted in a significant increase in blood utilization in 2010. These effects dissipated in 2011 and 2012, indicating that the disruption was strictly a short-run phenomenon. As expected, control groups such as cardiovascular surgery and urology experienced no statistically significant changes in mean blood use. These event-study results are illustrated in Figure 4.

**Figure 4:**
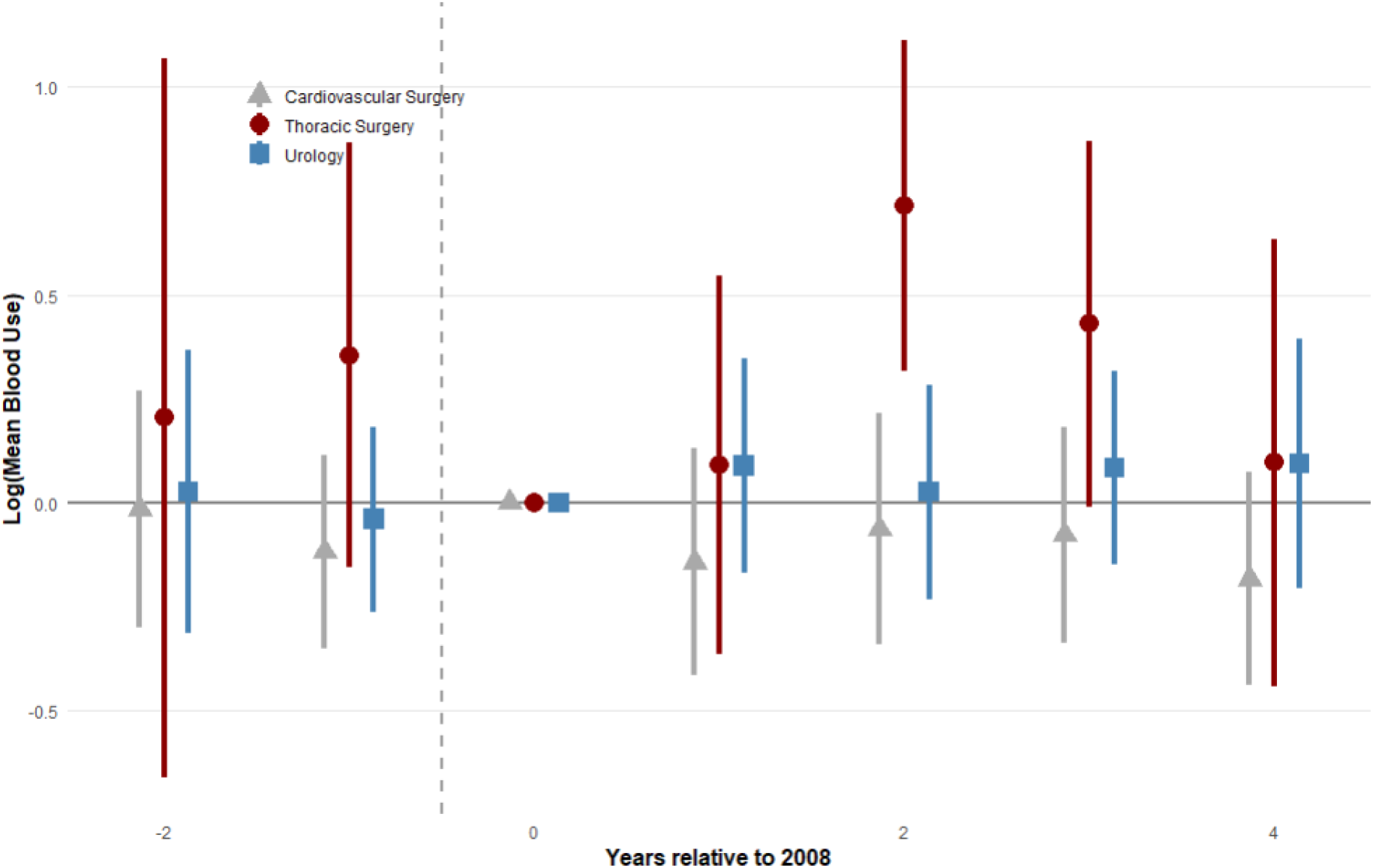
Effect of Regionalization on logged mean blood use (Index=0 is 2008)

The primary coefficient of interest for thoracic surgery in 2010 yields a log-difference of approximately 0.7144 . Converting this coefficient via *e*^0.7344^ − 1 ≈ 104.3%indicates that, relative to the reference year, mean blood use increased by roughly 104.3%among patients managed by thoracic surgeons. Because this marks an isolated significant surge within the dynamic framework, we implemented a secondary robustness which changes the composition of the control group. As the original estimation used all surgical specialities, a total of 7 different specifications is considered. The distribution of t-values for these results are displayed in Figure 5. It can be seen that regardless of the set of control specialties, the increase in of logged mean blood among thoracic surgeons remains significant.

**Figure 5:**
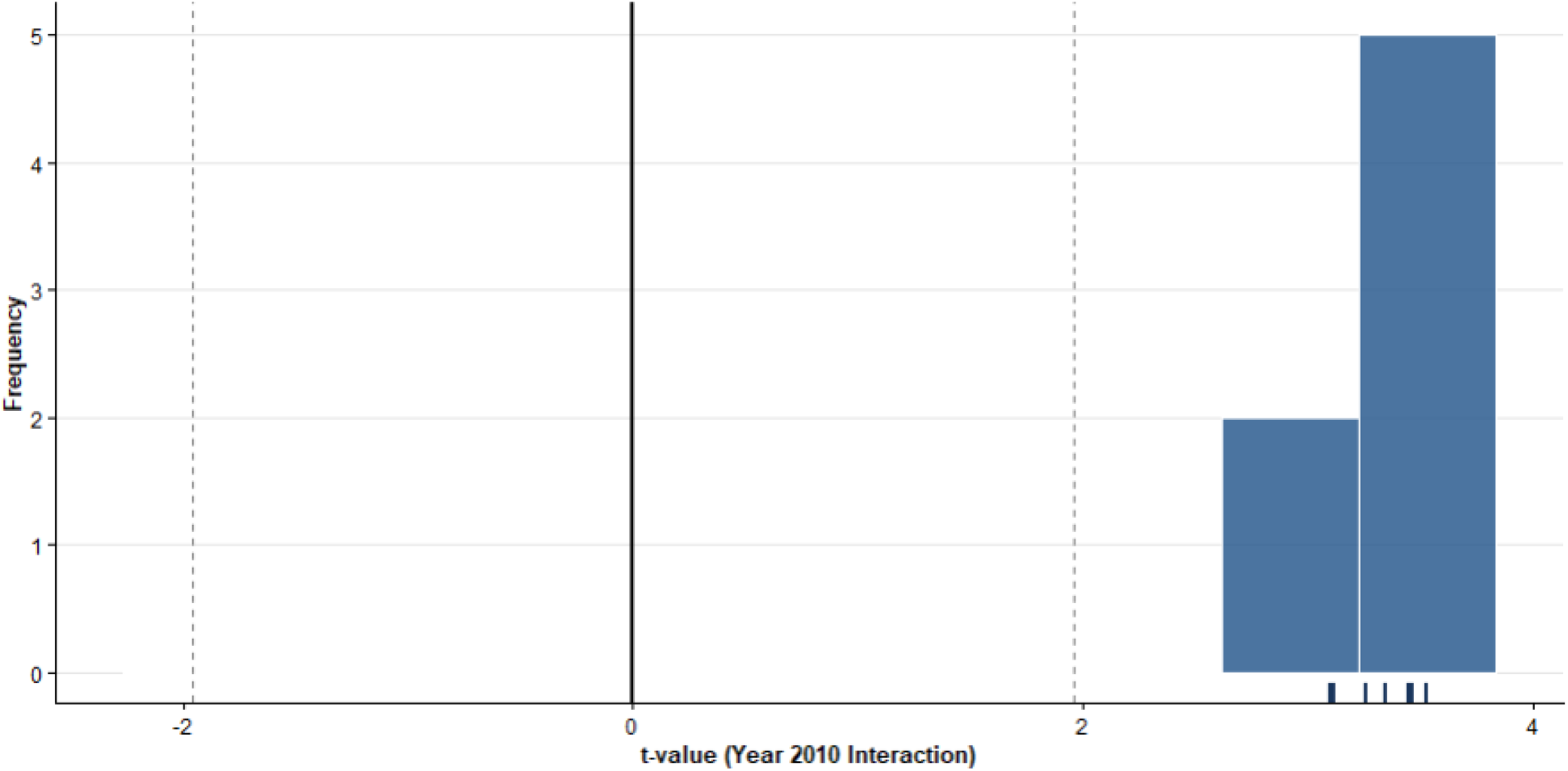
Distribution of t-values following permutation of control group.

## 5. Discussion and Limitations

This study investigates the unintended consequences of Cancer Care Ontario’s regionalization policy on blood product utilization, focusing on the centralization of thoracic oncology surgery at The Ottawa Hospital. Using a dynamic difference-in-differences framework paired with rigorous randomization inference, we document a sharp, statistically significant short-run surge in mean red blood cell transfusion rates among thoracic surgery patients following the opening of the new cancer centre in December 2009. Specifically, mean blood use increased by roughly 104.3% in 2010 before dissipating in subsequent years, while control specialties (cardiovascular surgery and urology) exhibited no parallel change.

These findings offer a novel perspective on how structural health policy interventions intersect with micro-level medical decision-making. Existing literature evaluating Ontario’s regionalization initiative has primarily concentrated on primary institutional objectives, noting that centralization successfully increased procedure volumes without immediately altering short-run operative mortality (Bendzsak et al., 2017). However, the results demonstrate that organizational shocks of this scale can induce significant secondary operational strains—specifically regarding blood resource utilization.

To interpret this Increase, we look to a theoretical mechanism established by a population-adapted treatment threshold model. Because the regionalization policy redistributed patient volumes and relocated community thoracic surgeons into a high-volume centre, it altered the distribution of physician beliefs and institutional risk tolerances. As smaller-community hospital surgeons—accustomed to different baseline patient risk profiles and variable complication rates—were integrated into the provincial center, their individual treatment thresholds shifted. In an environment where blood transfusion decisions are guided entirely by individual physician judgment rather than explicit financial incentives, this labor supply shock manifested as a temporary surge in precautionary or practice-variant blood utilization. The transient nature of the effect (fading by 2011–2012) further suggests that clinical practice patterns eventually re-converged as physicians adapted to the institutional standards of the new regional centre.

Despite its contributions, this study is subject to several limitations. First, due to data limitations stemming from aggregate secondary extraction (Shehata et al., 2014), granular, patient-level clinical characteristics such as preoperative hemoglobin levels, exact cancer staging, or intraoperative complications could not be controlled for. Second, while permuted specifications strongly support the robustness of the 2010 increase, the analysis relied on aggregate hospital-level proxies rather than direct physician-level tracking. Another important limitation was that the regionalization of thoracic surgery moved both surgeons and their patients. This creates an issue with identifying if this change was due to movement of more severely affected patients or if it was inclusive of beliefs. This backdoor path can only be closed if it is assumed that patient type is consistent across the province of Ontario. Future research leveraging micro-level administrative data could further isolate the exact behavioral pathways driving clinician-specific transfusion variability during this large-scale restructuring.

## 7. Conclusion

This paper examined how Cancer Care Ontario’s regionalization of lung cancer surgery impacted blood product utilization at The Ottawa Hospital. Combining a dynamic difference-in-differences framework with a population-adapted treatment threshold model, we linked hospital-level fluctuations in blood transfusions to micro-level clinical decision-making. Our findings indicate that the centralizing of specialized surgical care induces short-run operational frictions, as the influx of community surgeons into regional centers temporarily alters institutional treatment propensities and resource utilization patterns. Ultimately, this study highlights that structural health reforms carry hidden organizational costs extending beyond immediate patient outcomes. While regionalization successfully concentrates complex procedures in high-volume settings, policymakers must anticipate the behavioral and resource adjustments required during workforce transitions. Future research should leverage granular, patient-level administrative data to explore clinician-specific decision-making pathways and how institutional memory embodied by treatment norms can transfer following the regionalization of care.

## Data Availability

All data produced in the present study are available upon reasonable request to the authors.

## Acknowledgements

This paper was prepared for the Georgetown Blood and Plasma Research Group Fellowship at the Institute for the Study of Markets and Ethics. Jacob Smith would like to express thanks to Peter Jaworski and Bill English for their support throughout the fellowship.

## Appendix: Supplementary Figures

**Figure 1A:**
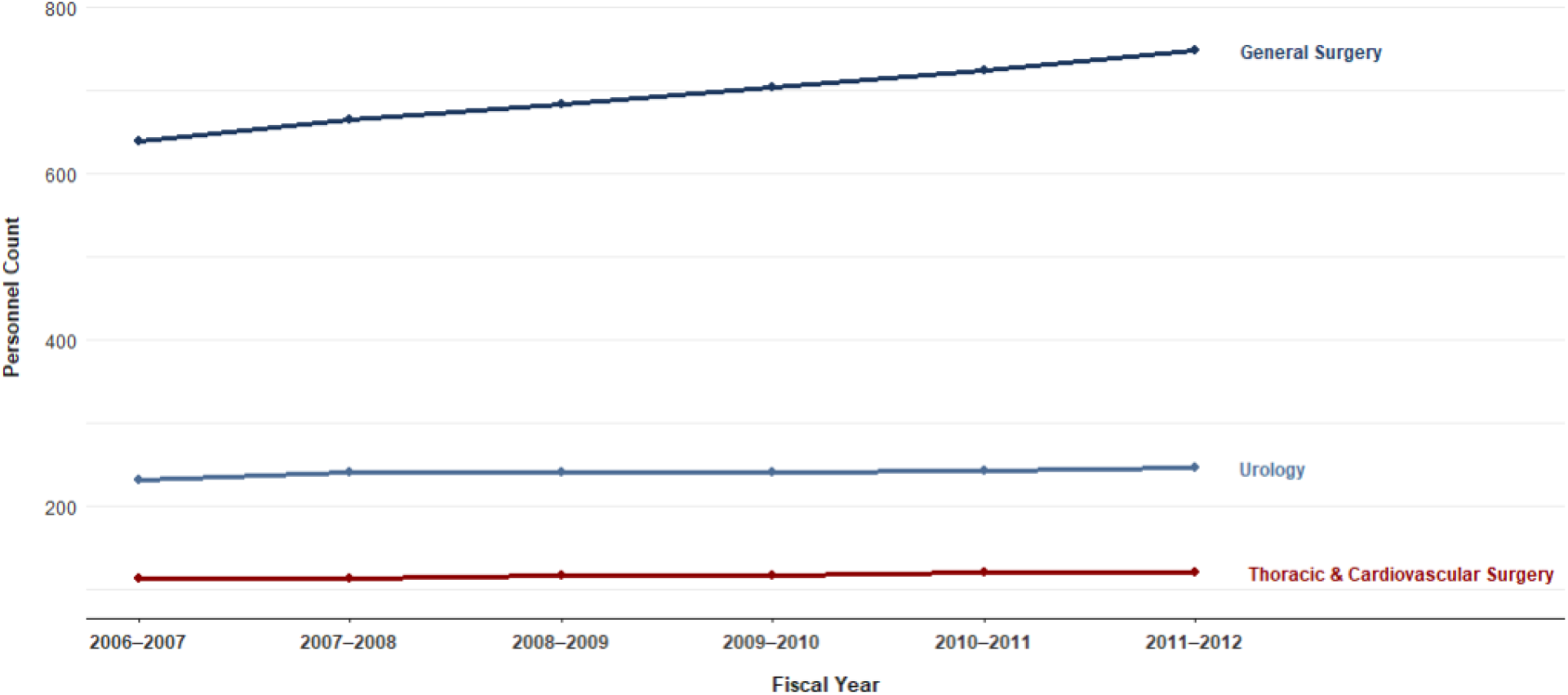
Provincial Level Personnel Counts across Surgical Specialties 2006-2012

